# Alice in Wonderland syndrome (AIWS): A research overview

**DOI:** 10.1101/2020.08.12.20173815

**Authors:** Md Mahbub Hossain

**Affiliations:** Texas A&M School of Public Health

**Keywords:** Alice in Wonderland syndrome, Todd’s syndrome, Pediatric Neurology, Neuroscience, Bibliometrics

## Abstract

**Background:** Alice in Wonderland syndrome (AIWS) is a rare neuropsychiatric condition characterized by distorted visual perceptions, body schema, and experience of time. This bibliometric study aimed to analyze the characteristics of the global research landscape on AIWS.

**Methods:** Bibliometric data on AIWS related publications published until 2019 were retrieved from the Web of Science database. The data were analyzed using statistical and scientometric tools to evaluate the publication trends, key research domains, top contributing journals, institutions, and countries associated with AIWS-related research.

**Results:** A total of 125 published items were analyzed with a mean of 3 authors and 8.15 citations per document. Most articles were published after 2008, in medical journals focused on neuropsychiatric sciences, and most institutions affiliated with AIWS research were based on high-income countries. Major research domains associated with AIWS included visual disturbances, body image, migraine, infections, risk factors, and other clinical correlates.

**Conclusion:** The current research landscape informs a developing trend in AIWS research in selected regions and specialties. Future research should emphasize multidisciplinary and translational investigations through global collaborations to advance the knowledge and practice on AIWS.

## Introduction

Alice in Wonderland syndrome (AIWS), also known as Todd’s syndrome, is a perceptual disorder, which is often characterized by impaired visual perception or metamorphopsias, abnormal body schema, and distorted experience of time [1–4]. The term “Alice in Wonderland syndrome” was adopted from the world-famous book titled “Alice’s Adventures in Wonderland” by Lewis Carroll, where the central character “Alice” perceived that the size and shape of her body changed in different scenarios [4]. In 1955, British psychiatrist John Todd (1914-1987) introduced the term “Alice in Wonderland syndrome” to describe a set of symptoms associated with migraine, epilepsy, and many other neuropsychiatric conditions [2]. Notable scholars, including Coleman and Lippman, made significant contributions in explaining AIWS [2, 4]. A systematic review identified 42 visual symptoms, 16 somesthetic, and other symptoms among AIWS cases [4]. These symptoms are commonly expressed as distortions of sensory perceptions rather than illusions or hallucinations. AIWS has been found to be associated with infection with pathogens such as Epstein Barr virus, migraine, depression, epilepsy, and delirium [3–5]. As is a rare neuropsychiatric condition, scientific knowledge on the etiology, epidemiology, clinical and psychosocial consequences associated with AIWS remains largely unexplored [2, 4, 6].

Recent methodological and technological advancements such as functional imaging techniques have resulted in a growing interest in AIWS and how the brain’s networks function among AIWS-affected individuals [4]. However, little is known about the research landscape of AIWS, which may offer valuable insights regarding future studies and scholarly discourses in this domain. Research overviews using scientometric or bibliometric methodologies are widely used to quantitatively evaluate the scientific publications in a domain or topic of research [7–10]. Also, such assessments inform the overall growth and trends of development in a field, thus prioritizing research agendas and mobilizing resources for addressing gaps in different branches of knowledge [11–13]. This study aimed to conduct a bibliometric evaluation of global publications on AIWS. Specifically, this study assessed the characteristics of scholarly articles, identified top contributors, and examined the key research domains in AIWS-related research.

## Materials and methods

In this study, bibliometric data were retrieved from Web of Science core collection using the following search query: *“Alice in Wonderland syndrome” OR “syndrome of Alice in Wonderland” OR “Todd’s syndrome”*. Web of Science core collection provides scholarly articles from biomedical and social sciences multiple databases, which has competitive bibliometric advantages over other databases [14]. After extracting the available literature, the titles and abstracts of those articles were assessed, and any citation relevant to AIWS irrespective of the document type, country of origin, and publication language were included in this study. This broad eligibility was adopted due to the lack of availability of studies on a rare condition like AIWS. All articles published from 1900 to 2019 were considered eligible for this study.

After screening the eligible studies, bibliometric analyses were conducted to examine the existing scientific landscape as studied in previous bibliometric studies [7, 14]. At first, a descriptive summary of major bibliometric characteristics was prepared, including the frequency distribution of published studies, their sources, indexed keywords, author appearance, collaboration, and citation index. Moreover, the top five journals, countries, and institutions were identified using the number of published documents.

Furthermore, major research domains related to AIWS were identified using the co-occurrence of keywords appearing more than once, which provided a visualization of connected topics using a mapping approach. Such mapping is prepared through a text mining-based network analysis approach as used in previous bibliometric studies. Each domain is mapped using different colors that examine the common topics represented in a research area and how multiple areas are related to each other. In this bibliometric study, reference management, descriptive analyses, and keywords mapping were conducted using *RefWorks*, *R* (Version 3.6.1), and *VOSViewer* software, respectively. This bibliometric study was registered in the Open Science Framework [15].

## Results

### An overview of existing studies on AIWS

A total of 125 articles were identified that met the criteria for this study (summary provided in Table 1). Most of these articles were original articles (n = 64) followed by reviews (n = 14), correspondence (n = 15), editorials (n = 11), and other types of publications (n = 23). These articles were published in 83 scholarly sources and had 178 keywords. There were 375 authors with a mean of 3 authors per documents and a collaboration index of 3.46. The mean citation per document was 8.15.

**Table 1:** Summary of the bibliometric findings.

| *Description* | *Results (frequency)* |
| --- | --- |
| **a. Overview of existing literature** |  |
| Total published studies (1977-2019) | 125 |
| Sources (Journals, reports, books, etc.) | 83 |
| Authors' keywords | 178 |
| Total authors | 375 |
| Authors of multi-author documents | 356 |
| Authors per document | 3 |
| Co-authors per document | 3.43 |
| Collaboration index | 3.46 |
| Mean citations per document | 8.15 |
| **b. Types of documents** |  |
| Original articles | 62 |
| Review articles | 14 |
| Letters/correspondence | 15 |
| Editorials | 11 |
| Others | 23 |

### Global publication trends on AIWS

The earliest publication on AIWS indexed in the Web of Science database was published in 1977. The number of publications per year remained low until 2008 (Figure 1), which started increasing afterward, and the number of publications reached a pick in 2018. The global map showing country affiliations associated with the available publications shows that countries in North America and Europe were affiliated with more publications, whereas fewer studies were found from countries in Asia, Africa, Oceania, and South America (Figure 2). Specifically, the U.S. had the highest number of publications (n = 27) followed by Japan (n = 11), Italy and Germany (n = 9 each), and Spain and France (n = 8 each) as shown in Table 2. Moreover, top journals and affiliated institutions were identified using the frequency of published articles on AIWS. Pediatric Neurology published 4.8% (n = 6) articles, whereas all top journals publishing AIWS-related articles belonged to neuropsychiatric specialties. Furthermore, the Sapienza University of Rome was affiliated with the highest (9.6%, n = 12) publications among global institutions. All top affiliated institutions were in high-income countries.

**Figure 1:**
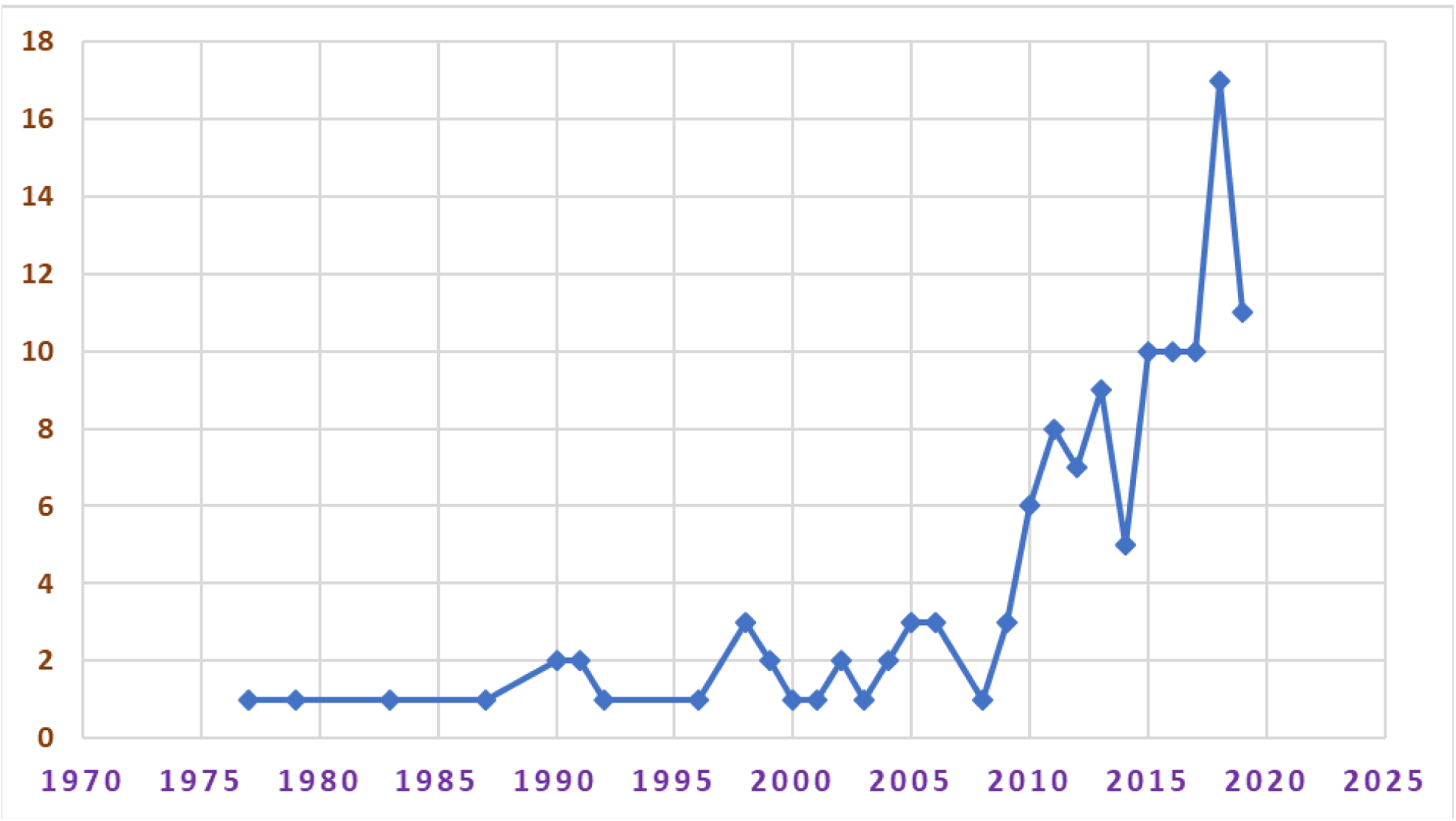
Trends of scientific publications on Alice in Wonderland syndrome.

**Figure 2:**
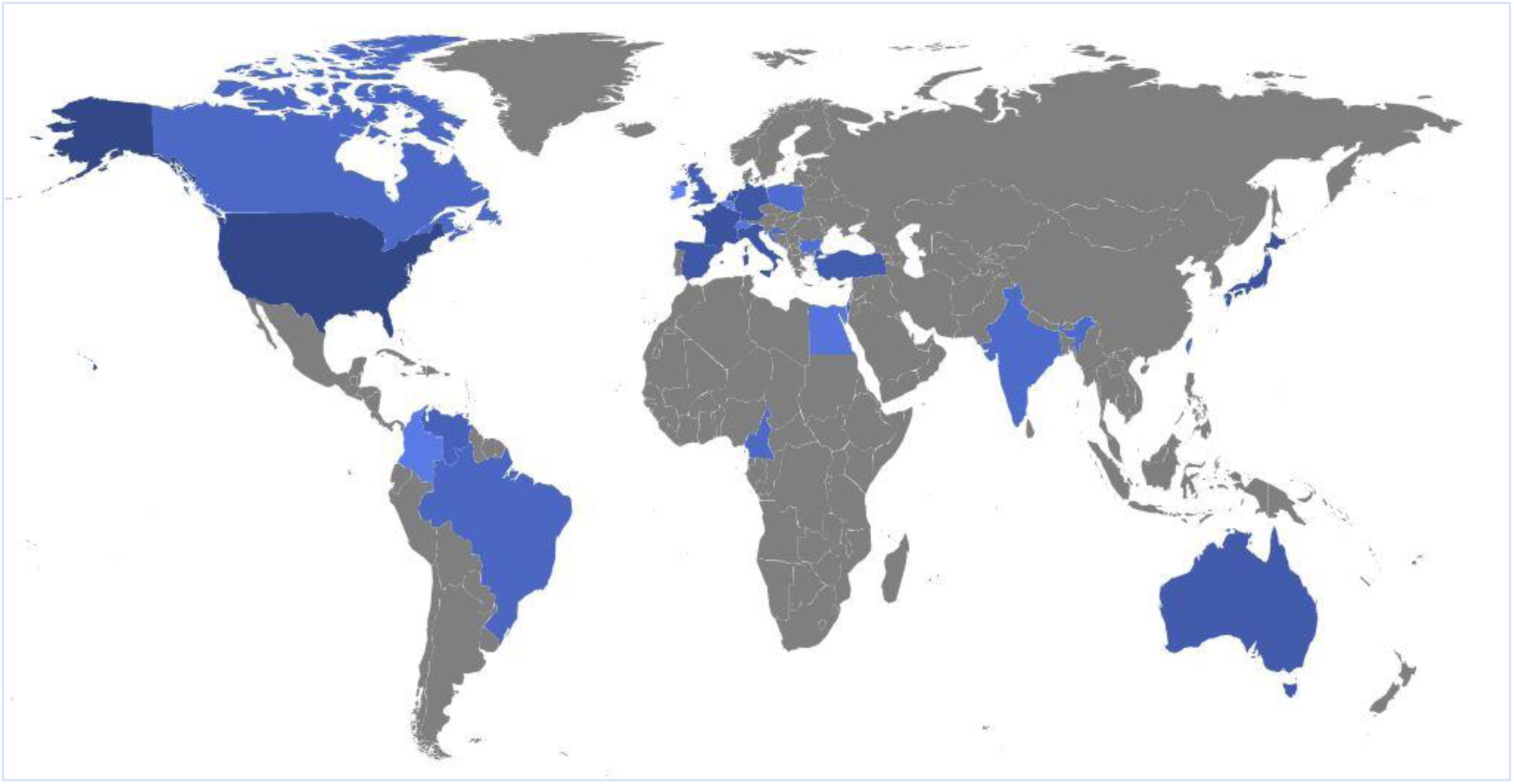
Countries that had contributed to research on Alice in Wonderland syndrome. (blue areas indicate countries contributing to AIWS research were marked whereas grey areas represent countries with no publication in this topic)

**Table 2:**
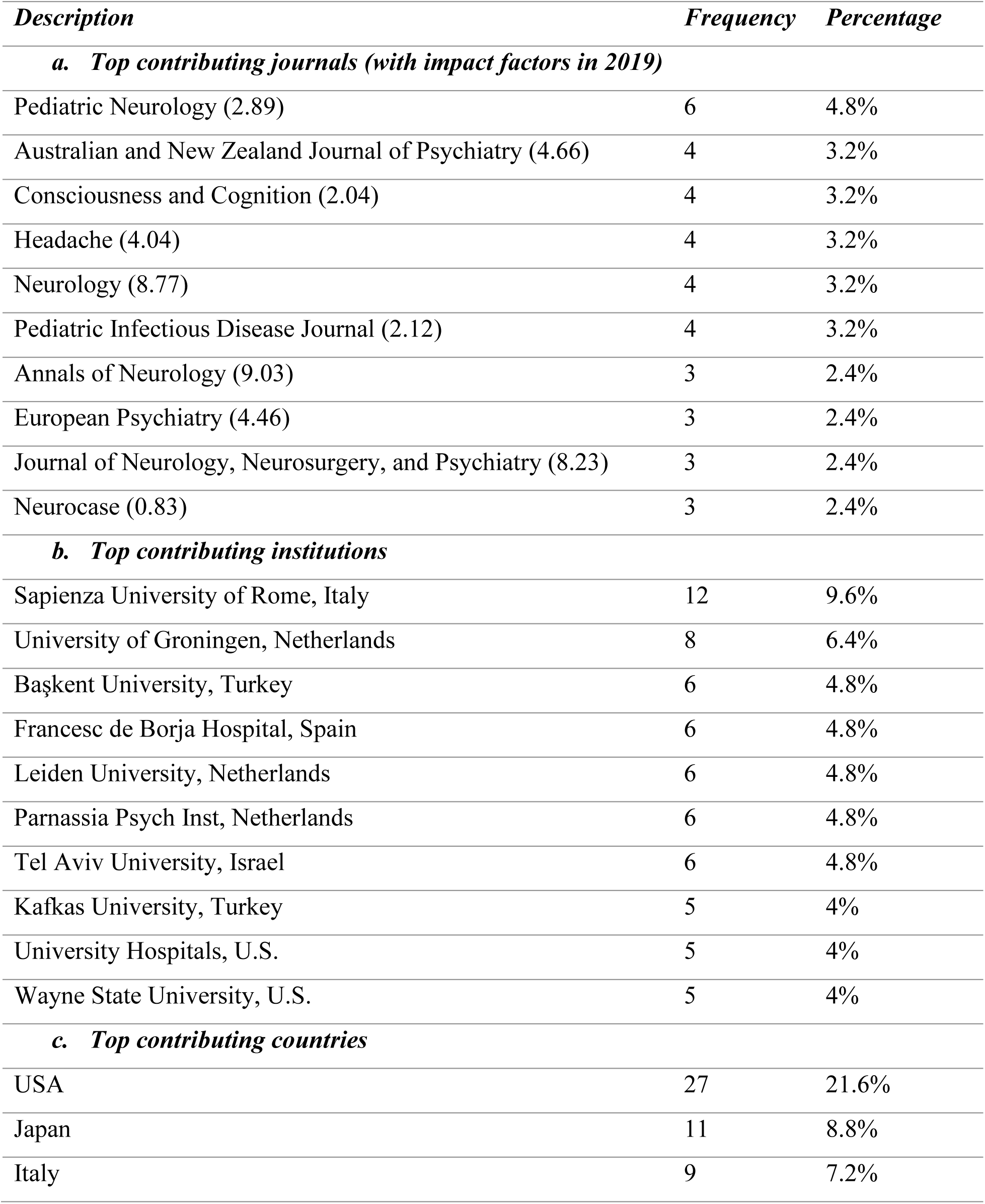
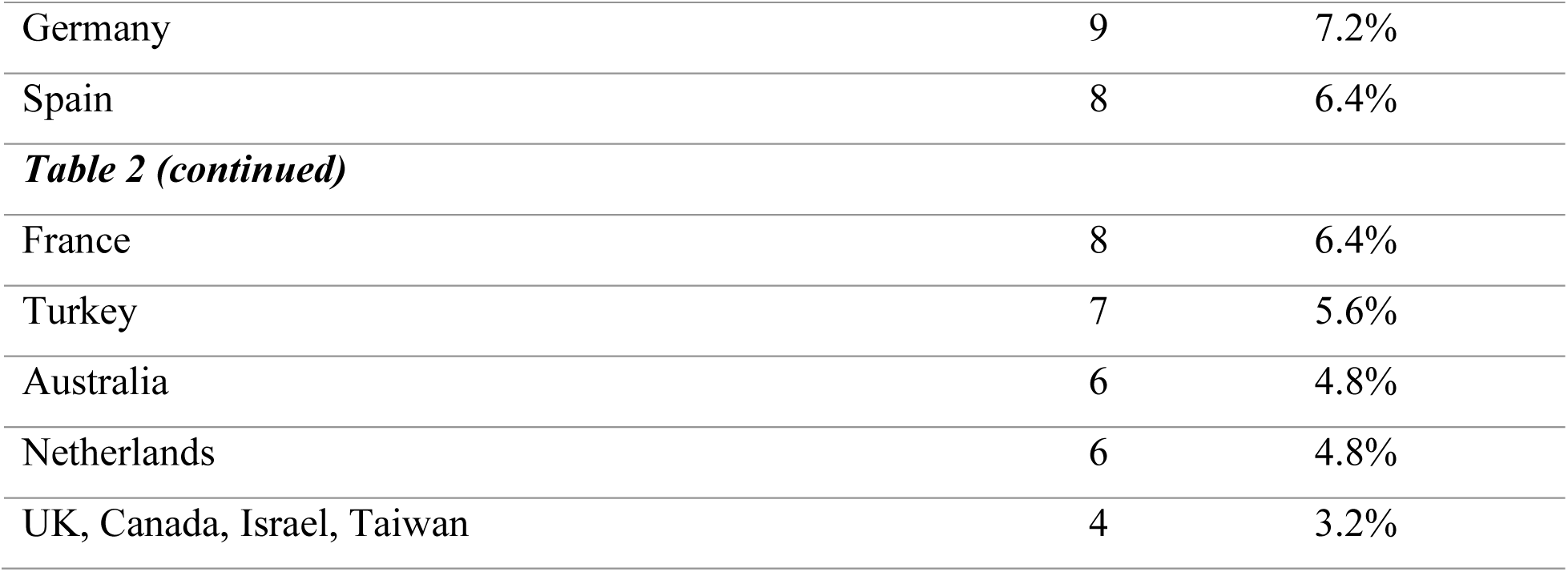
Top contributing journals, affiliated institutions, and countries.

### Research domains associated with AIWS

Several research foci were identified where multiple keywords clustered, highlighting the cooccurrence of those topics across the literature. One such domain is related to clinical features associated with AIWS that are often reported by the patients, including altered perceptions regarding body image, schema, and anorexia nervosa, as highlighted in yellow color in Figure 3. For example, Pitron and colleagues described conceptual factors highlighting that the body schema has some primacy over the body image acknowledging the special role played by the body image in neuropsychiatric conditions such as AIWS [16]. Another study by Pozo and colleagues reported that 45% of the individuals with AIWS had disturbances in body image alongside other clinical features [17].

**Figure 3:**
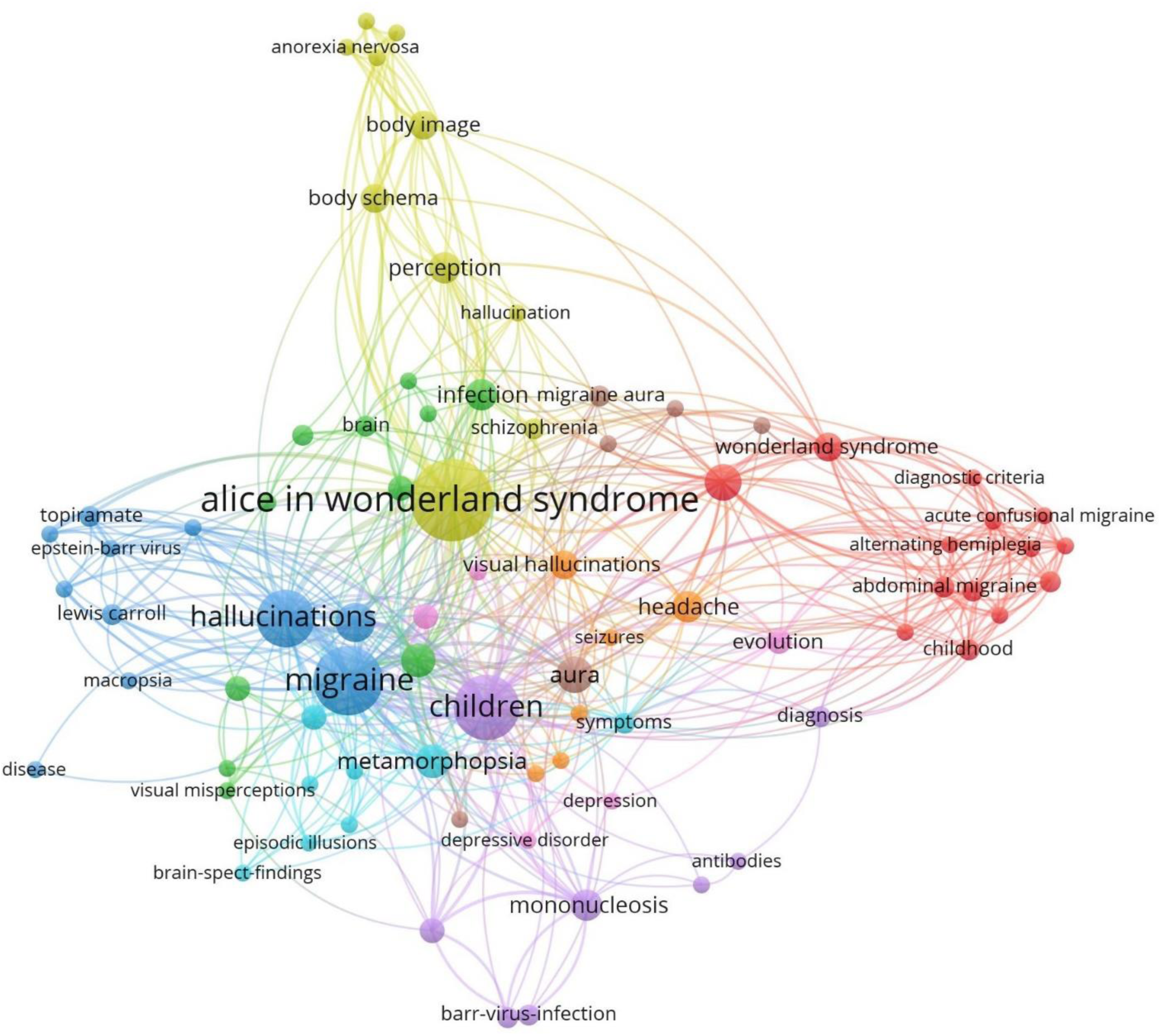
Mapping keywords associated with Alice in Wonderland syndrome (AIWS)

Two more domains were found, which emphasized on migraine and associated clinical conditions as highlighted in blue and red in Figure 3. The complexity of migraine subtypes and cooccurrence with multiple neurological conditions have been reported in the existing literate. For example, a case of abdominal colic was found to have occurred with AIWS [18]. Another study reported that nearly 77% of AIWS occurred during the vestibular migraine [19].

Furthermore, a major proportion of research indicates childhood infections as a research area within AIWS literature, as shown in purple in Figure 3. Infection with Epstein-Barr virus or mononucleosis was repeatedly found to be associated with AIWS [20–24]. For example, Cinbis and Aysun reported a case of visual metamorphopsia alongside the course of infectious mononucleosis [20]. Multiple case reports had shown the occurrence of AIWS in infections diseases such as Coxsackie B1 virus encephalitis, Influenza A virus encephalitis, Cytomegalovirus, Varicella-zoster encephalitis; lesions in the central nervous system including Acute disseminated encephalomyelitis, Cavernous angioma, Traumatic encephalopathy, and Wallenberg syndrome; and neuropsychiatric conditions including epilepsy, depression, schizophrenia, and misidentification syndrome [4]. Furthermore, common keywords related to symptoms and diagnostic features appeared between multiple domains highlighting potential cooccurrence and overlaps between research themes within the umbrella of AIWS.

## Discussion

This bibliometric overview of the current research landscape shows a limited number of articles published on AIWS. Most of the articles were published in neuropsychiatric journals, and most authors were affiliated with institutions located in high-income countries in Europe and North America. A lack of primary studies and a poor representation of low-income countries provide several insights on the research trends in the realm of AIWS. One possible explanation for these trends is the rarity of AIWS in the general population [4, 25, 26], which may have contributed to a low number of studies globally with even a lower number of studies from low-income countries. Another challenge can be a lack of research capacities in those countries that share a major proportion of the global burden of diseases yet have fewer publications on critical health problems [27–29]. Moreover, as AIWS is mostly studied in children within clinical settings [4], community and population-based studies of AIWS, associated factors, prognosis, and outcomes are yet to be studied. However, academic and professional interests on AIWS is increasing in recent years, as reflected in the publication trends. Advanced research on AIWS and clinical correlates should be promoted globally using standardized tools and techniques. The ontology and phenomenology of AIWS should be investigated through organized global research, which may minimize the errors in creating and synthesizing evidence in this domain.

The current literature highlights an association between infectious diseases at an early age and neuropsychiatric manifestations associated with AIWS [3, 4]. Such complexities in the pathophysiological, neurological, and psychiatric features make AIWS an appealing case for the healthcare providers and researchers from multiple domains, including microbiology, immunology, child health, neurosciences, behavioral sciences, and mental health sciences. Multispecialty research investigations across population sub-groups may provide epidemiological insights on AIWS and help understanding how the human body and mind interact at the edge of multiple risk factors and manifest clinical features among the affected individuals [30–32]. Nonetheless, a deeper understanding of AIWS can facilitate scientific discourses on multiple pathways involved in body systems, especially when someone is infected with viruses or lesions that are associated with AIWS.

The growth of scientific literature in a topic not only reflects the interests of the academic community but also provides a basis to make clinical decisions as well as health policies. Rare diseases that are inadequately studied, often underrecognized or lately diagnosed, and those disorders may leave the patients and their caregivers helpless if the healthcare ecosystem does not offer effective treatments at affordable costs [33]. The recent developments in rare diseases and orphan drugs inform the need for studying biomedical conditions that affect individuals and families. It is essential to consider these issues while promoting academic and translational research on AIWS and associated clinical conditions for improving patient-centered care.

Furthermore, the available research necessitates a cross-disciplinary exchange of research evidence on AIWS to promote preventive and therapeutic strategies. Such efforts in translating research into clinical and public health practice may encourage the scientific community to conduct further research and enrich the evidence base on AIWS. As AIWS is a rare clinical condition, there is a need for collaborations among physicians and researchers across disciplines as well as among institutions around the world.

This study has several limitations that should be acknowledged. Although Web of Science is one of the major bibliometric databases that provide extensive data on scholarly publications, there are other databases that could provide a different set of insights compared to the current study. The choice of database and methodological approaches to the bibliometric data can induce selection bias and affect the generalizability of this study. Another major concern in a potential publication bias as not all cases or studies on AIWS are published in indexed journals. Such studies, if there are any, could have provided further scholarly perspectives on the existing research landscape. Such limitations should be addressed in future research and intellectual discourses on AIWS.

## Conclusions

This study assessed global research publications on AIWS using a bibliometric approach. The current publication trends suggest a growing interest in AIWS in recent years, whereas a limited number of studies from selected regions inform the need for extensive research collaborations exploring diverse areas associated with AIWS. In addition, multidisciplinary research should be encouraged at the institutional level to understand the neuropsychiatric pathophysiology of AIWS across populations. The existing literature highlights the importance of extending research efforts and translating available evidence for healthcare decision-making. It is essential to encourage and enable scholars and practitioners to leverage collaborative skills and strengthen collective measures to understand and prevent AIWS and associated adverse health outcomes.

## Data Availability

This study is registered with the Open Science Framework (OSF). The underlying data associated with this study can be accessed here:
Hossain MM. Global research overview of Alice in Wonderland syndrome (AIWS): a bibliometric study. doi:10.17605/OSF.IO/GXWFP.

https://osf.io/7d9vn/

## Acknowledgments

None.

## Conflicts of interest

None.

## Funding statement

No funding was received at any stage of conducting this research.

## Notes

### Competing Interest Statement

The authors have declared no competing interest.

### Author Declarations

This is a bibliometric study that did not involve any human participants. Therefore, no IRB approval was required for this study.

